# Appraised evidence and decision making in clinical practice

**DOI:** 10.1101/2024.11.15.24317373

**Authors:** Chrysi Leliopoulou, Theodora Stroumpouki, Emmanouil Stafylarakis, Linda Collins

## Abstract

**Background:** The use of evidence in decision making in clinical practice may vary significantly from practice to practice. Research suggests that clinicians may ignore research-based evidence in their practice or use **research** evidence indirectly. The aim of this systematic review is to explore using an online independent learning approach for clinicians making evidence-based decisions using research and evidence.

**Methodology:** This systematic literature review was conducted across MEDLINE, PUBMED, SCIENCE Direct, APA PSYCINFO and CINHAL databases. The use of a search strategy employed to conduct searches on the individual database. Using the MMAT criteria by Hong et al., 2018) studies were screened and reviewed.

**Results:** The findings from each study were thematically analysed and organised in the following thematic areas: (1) Searching for an answer to the clinical question or problem; (2) Expanding on the evidence to address the clinical question or problem; (3) Recording the available evidence to the clinical problem; (4) Validating the evidence, whether research-based or tacit knowledge; (5) Inspecting the evidence for its potential to improve current practice; (6) Cascading the evidence appropriately and possibly adopting the evidence in the decision; and (7) Evaluating the outcome of the clinical decision.

**Conclusions:** This systematic literature review found there is a lack of an online independent learning approach for clinicians making evidence-based decision using research and evidence which highlights a gap in the literature.

## Introduction

Health professionals are urged to appraise research-based evidence and inform their decision-making processes in clinical practice to safeguard the delivery of effective and person-centred care (Mai et al., 20221; Kislov et al., 2019; Rycroft-Malone, 2004). Health professionals would need to consider the quality of clinical guidance and its relevance to their practice, but also professionals are required to appraise new and complex research based evidence and implement this in their clinical practice (Higgs and Titchen 2000).

Tools such as the Grading of Recommendations Assessment, Development, and Evaluation (GRADE, 2023) and the Appraisal of Guidelines for Research and Evaluation (AGREE, 2017) support professionals access evidence easier and integrate such evidence to improve their decision making and patient experience. Such tools, however, can be time consuming, complex, and expensive to implement (Al-Jundi and Sakka, 2017; Twells, 2015; Friesen-Storms et al., 2015) which may explain why adoption of research-based evidence remains low in clinical practice (Kislov et al., 2019).

The aim of this systematic review is to identify how online independent learning may inform and transform clinical decisions utilising evidence-based practice. This review seeks to encourage frontline clinicians to integrate research and evidence into their daily decision-making processes as part of online independent learning.

## Methodology

### Search Strategy and Selection Process

Four researchers conducted a literature search using the following strategy: (screening evidence OR decision-making OR clinical practice inquiry) AND (nursing practice) AND (critical thinking OR clinical decision). This search strategy was used to search CINHAL, MEDLINE, SCIENCE Direct, APA PSYCINFO and PubMed databases. Additionally, a manual search of reference lists from all selected articles was performed to identify any studies that may have been missed.

The search strategy developed using keywords and Medical Subject Headings (MeSH), targeted terms related to evidence-based practice, critical appraisal, and evidence screening. Synonyms and the Boolean operator “OR” were used to search for terms simultaneously, while “AND” was used to link related topics within article titles or abstracts (Power and Siddall, 2015). The search included terms such as critical appraisal, screening evidence, evidence-based practice, decision-making, clinical practice inquiry, and health professionals. The researchers also manually scanned reference lists of selected articles to ensure no relevant studies were overlooked. The databases searched can be found on figure 1 which reports the number of records identified from each database searched and included after inclusion criteria applied.

**Figure 1:**
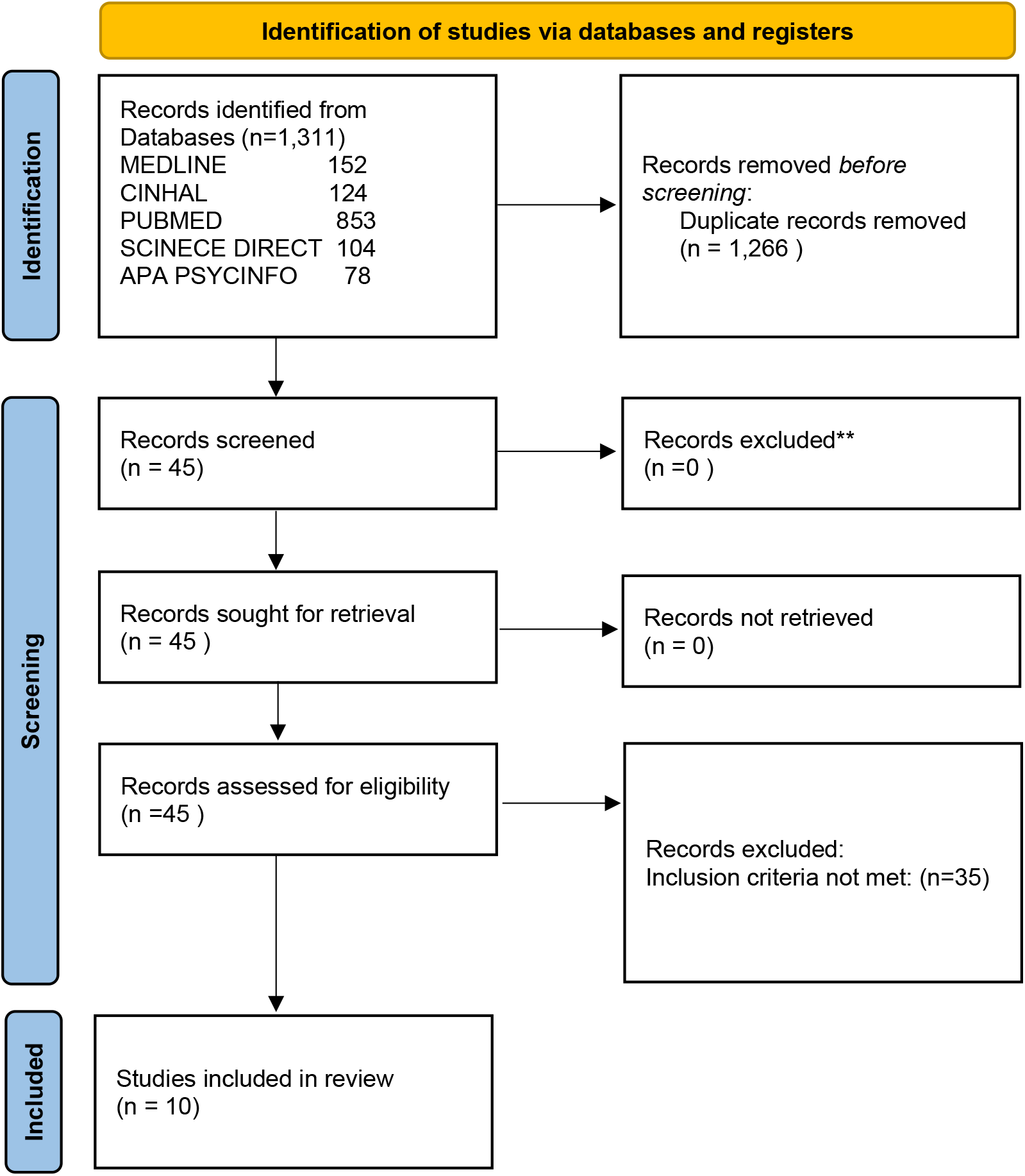
This figure shows how studies were screened using the PRISMA guidelines.

The selection process followed a two-step approach, in line with the recommendations of Polanin et al (2019) and Aromataris and Pearson (2014). Studies published between 2014 and 2024, written in English, and targeting nurses or allied health professionals were included. Researchers independently screened titles and abstracts, and full papers were retrieved for further review. From the final electronic searches, a total of 1311 articles were retrieved 239 and from those only ten met the MMAT criteria applied to screen the included studies (Hong et al., 2018).

Papers were reviewed following the Preferred Reporting Items for Systematic Reviews and Meta-Analyses (PRISMA) guidelines. After removing duplicates, the abstracts of forty-five articles were assessed, with thirty-five not meeting the inclusion and/or MMAT criteria, leaving ten full-text articles for further analysis and discussion.

### Data Analysis

This literature review found that there is a notable shortage of publications focusing on online independent learning health care clinicians may engage in order to retrieve the relevant evidence to inform their decision making. While frameworks and tools exist to assess research evidence, there is a lack of resources designed to help health professionals evaluate the quality of appraised research evidence incorporated in their decision making in clinical practice.

**Table 1:**
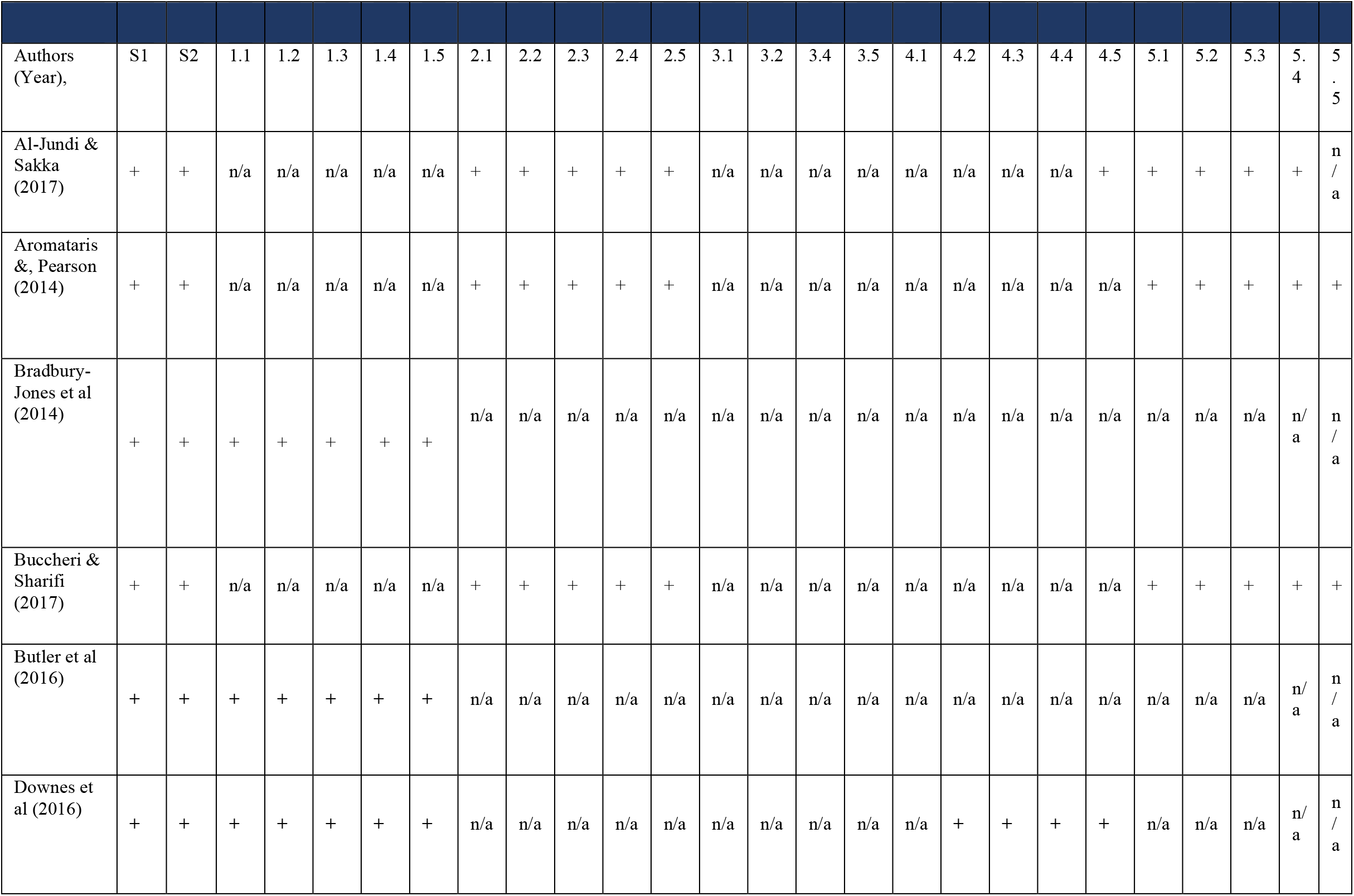
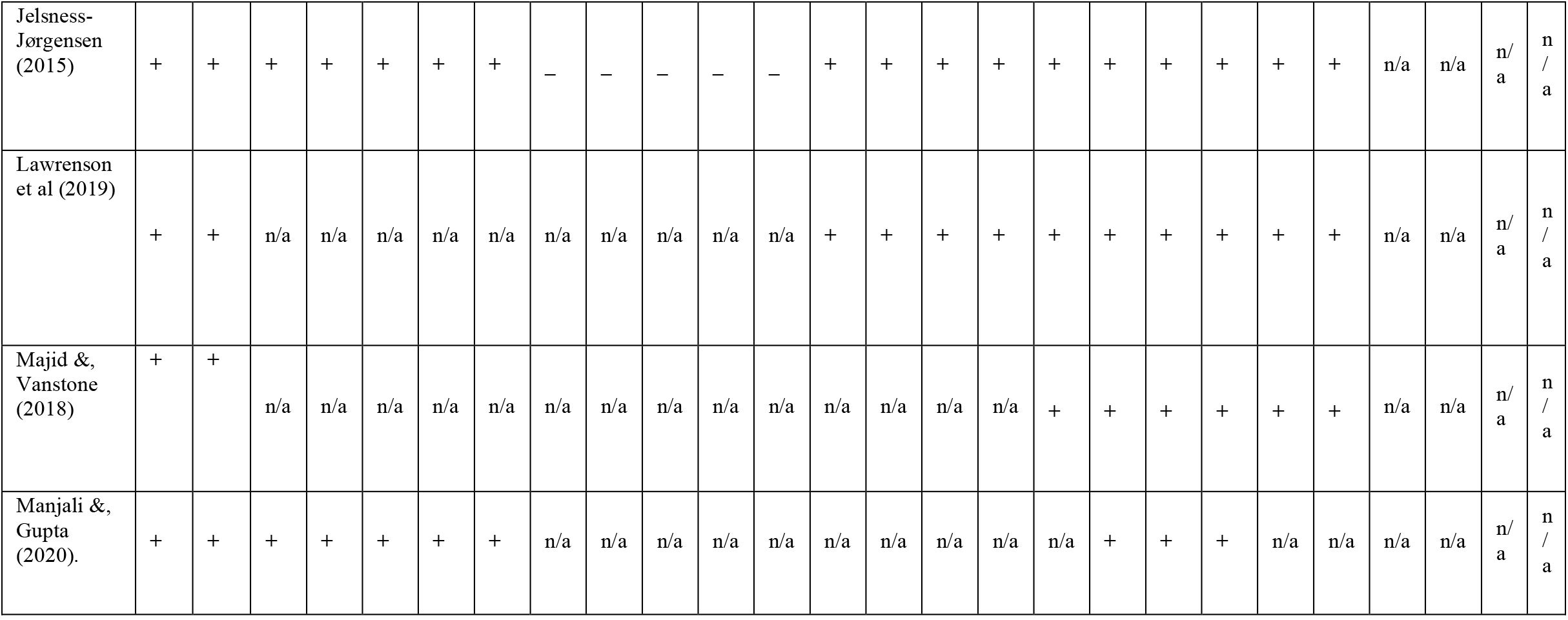
The use of MMAT appraisal tool (Hong et al 2018) was used to appraise the studies included.

**Table 2:** This table explains the MMAT quality criteria.

| Category of study designs | Methodological criteria |
| --- | --- |
|  | S1. Are there clear research criteria? |
|  | S2. Do the collected data allow to address the research question? a |
| Further appraisal may not be feasible or appropriate when the answer is “No” or “ can’t tell” to one or both screening questions |  |
| Qualitative | 1.1 is the qualitative approach appropriate to answer the research question? a |
|  | 1.2 Are the qualitative data collection methods adequate to address the research question? |
|  | 1.3 Are the findings adequately derived from the data? |
|  | Is the interpretation of results sufficiently substantiated by data? |
|  | 1.5 is the coherence between qualitative data sources, collection, analysis and interpretation? |
| Quantitative/RCTs | 2.1 is randomisation appropriately performed2.2. are the groups comparable at baseline?? |
|  | 2.3 are there complete outcome data? b |
|  | 2.4 Are outcomes assessors blinded to the intervention provided? |
|  | 2.5 Did the participants adhere to the assigned intervention? |
| Quantitative/Non-Randomised | 3.2 Are the participants representative of the target population? |
|  | 3.3. Are measurements appropriate regarding both the outcome and exposure/intervention? |
|  | 3.4 Are the confounders accounted for in the design and analysis? |
|  | 3.5 During the study period, is the intervention/exposure administered as intended? |
| Quantitative/Descriptive | 4.1 is the sampling strategy relevant to address the research question? A |
|  | 4.2 is the sample representative of the target population? |
|  | 4.3 Are the measurements appropriate? |
|  | 4.4 is the risk if nonresponse bias low? |
|  | 4.5 is the statistical analysis appropriate to answer the research question? |
| Mixed Methods | 5.1 is there an adequate rationale for using a mixed methods design to address the research question? a |
|  | 5.2 Are the different components of the study effectively integrated to answer the research question? a |
|  | 5,3 Are divergences and inconsistencies between quantitative and qualitative results adequately addressed? |
|  | 5.5 Do the different components of the study adhere to the quality criteria of each tradition of the methods involved?? |
| Total percentage quality criteria met |  |
| a +=yes, -=no, ?=can't tell, n/a=not applicable<br>b acceptable complete data value ranges from 80% to 95% (Higgins et al 2016; Thomas et al 2004; Zaza et al 2000) |  |

Interestingly, there is a clear lack of how clinicians independently learn to evaluate evidence in making evidence-based decisions using research and evidence. The studies included in this systematic review reveal that the appraisal process and the decision-making process share some key areas of consideration clinicians often evaluate when they are appraising research-based evidence and/or other types of evidence. In clinical practice, many decisions are made on the preferences of patients or available resources, thus professionals such as nurses may need to independently learn to refer to other types of evidence before they make an informed and clinically relevant decision.

## Results

This systematic literature review found that there are seven areas clinicians may concern themselves with when appraising any type of evidence either research-based evidence or local data. These areas may also inform their decision in clinical practice. These areas were drawn directly from the findings of the articles reviewed and they are as follows: (1) Searching for an answer to the clinical question or problem was a common starting point in the appraisal process of the information at hand; (2) Expanding on the evidence to address the clinical question or problem seemed important in order to understand the depth of the evidence available; (3) Recording the available evidence relevant to the clinical problem; (4) Validating the evidence, whether research-based or tacit knowledge in real life scenarios is important; (5) Inspecting the evidence for its potential to improve current practice is also critical; (6) Cascading the evidence appropriately in adopting the evidence in the decision is also crucial in order to have a team approach to the problem; and (7) Evaluating the outcome of the clinical decision is also an integral part of this.

These seven areas inform the acronym “S.E.R.V.I.C.E.” as a short reference to the process decision makers may resort when seeking evidence to inform their clinical decisions.

**Table 3:**
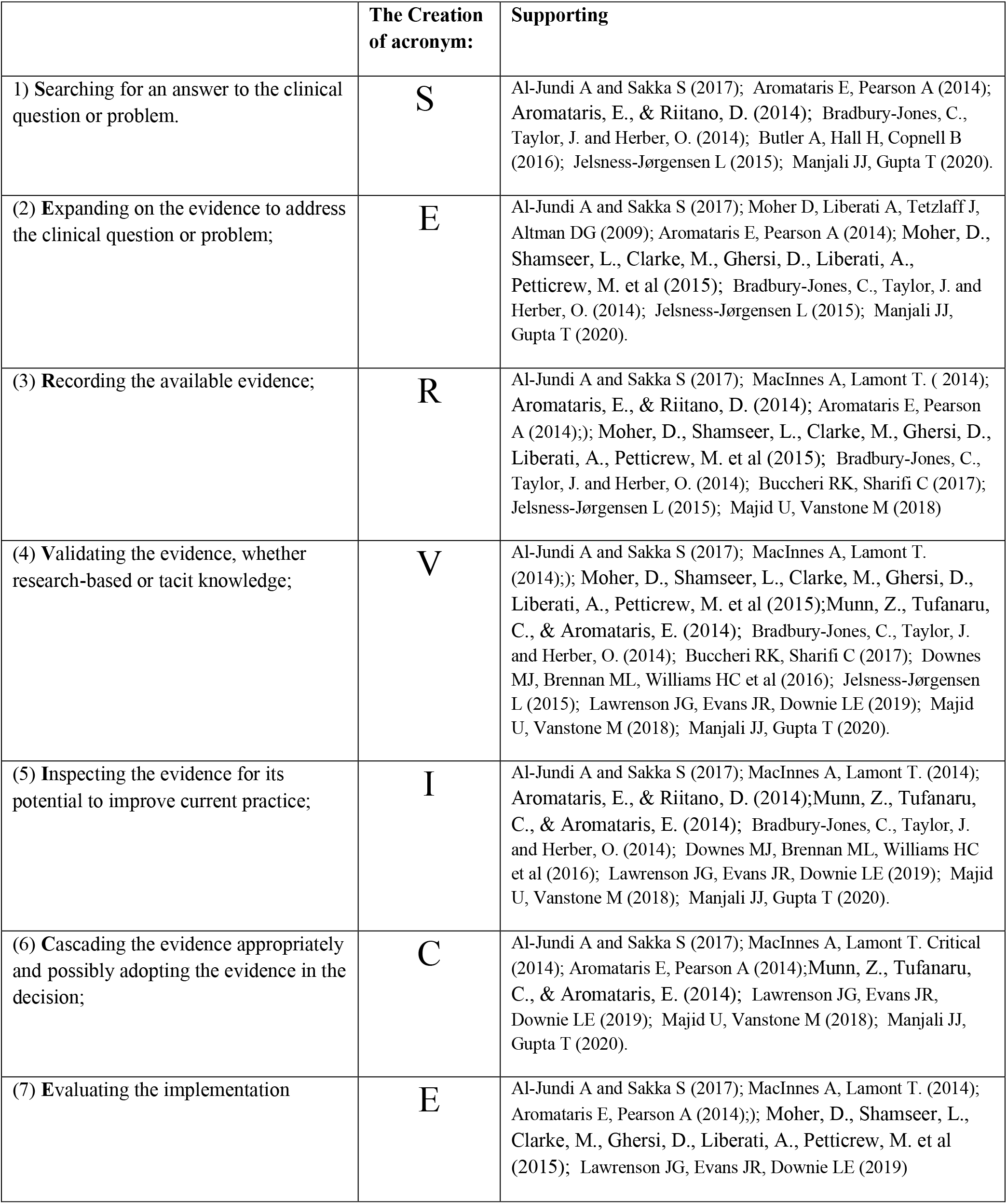
This table explains the creation of the acronym S.E.R.V.I.C.E.

## Discussion

This systematic literature review found that clinicians seek out evidence based answers to clinical questions or clinical problems regardless of the type of evidence (research-based evidence or tacit evidence). The seven areas of the acronym “S.E.R.V.I.C.E.” may help clinicians integrate evidence better in their decision making and may engage with evidence based practice more because the acronym is easy to use, it doesn’t take up a lot of clinical time and provides the clinician with some assurances around the quality of the evidence available to the clinicians in practice.

However, this review acknowledges the limitation of this review such as the small sample size of studies included, and the need to run a feasibility study to confirm the validity and reliability of the S.E.R.V.I.C.E. acronym (Baethge et al., 2019; Brouwers et al., 2010b).

The acronym was designed to be simple and user-friendly, with the potential to improve the overall quality of decision-making for busy clinicians and create an inclusive solution by ensuring that all forms of evidence, including tacit knowledge, are considered in decision making. This acronym may be used as part of an open independent approach to learning how clinical decisions can be made and promote an inclusive approach to evidence-based practice. This in turn may promote further engagement of clinicians with research based evidence practice, but it is also recommended that a larger-scale systematic review across various clinical settings may increase the acronym’s transferability.

## Data Availability

All data produced in the present work are contained in the manuscript.

